# Cardiovascular and Cardiometabolic Outcomes in Adults with Fetal Alcohol Spectrum Disorders: A Retrospective Cohort Study

**DOI:** 10.64898/2026.03.19.26348806

**Authors:** Olivia Weeks, Lynn A. Sleeper, Shaan Khurshid, Wolfram Goessling, C. Geoffrey Burns, Caroline E. Burns, CIFASD

## Abstract

**Background:** Fetal alcohol spectrum disorders (FASDs) impact up to 5% of U.S. school age children; however, the burden of cardiovascular disease (CVD) in adults with FASDs remains poorly defined. We investigated associations between FASDs, cardiometabolic abnormalities, and CVD using electronic health record (EHR) data.

**Methods:** We performed a retrospective cohort study of adults (≥18 years) receiving ambulatory care with a FASD diagnosis (*n* = 208, mean age 38.4±14.5, 50% female) and age- and sex-matched control patients without FASD (*n* = 824, mean age 41.6±14.5, 50% female). Cardiometabolic outcomes were overweight/obesity, dyslipidemia, and diabetes mellitus. Cardiovascular outcomes were congenital heart defects (CHDs), heart murmur, hypertension, conduction defects, arrhythmias, structural heart remodeling, systolic and diastolic dysfunction, heart failure, myocardial infarction (MI), stroke, and thromboembolic events. Associations were assessed using age-adjusted logistic and Poisson regression, with sex-by-diagnosis interaction testing. Multivariable logistic regression was then used to estimate the odds of cardiovascular outcomes with additional adjustment for cardiometabolic conditions. Associations between CHD and CVD outcomes were evaluated using Fisher’s exact tests.

**Results:** Adults with FASDs had a higher prevalence (p<0.05) of cardiometabolic abnormalities, including dyslipidemia, type 2 diabetes mellitus (T2DM), and the co-occurrence of multiple of these conditions (overweight/obese, HDL cholesterol < 40 mg/dL, and T2DM). CHDs were significantly more common in individuals with FASDs than controls (6% vs 1%, p < 0.001). Similarly, the FASD cohort had a higher incidence of systolic and diastolic dysfunction (6% vs. 2%), structural heart remodeling (11% vs 5%), MI (6% vs. 2%), stroke (4% vs 1%), and thromboembolic events (4% vs 1%; all p < 0.05). Significant sex-by-diagnosis interactions were observed for hypertension, arrhythmia, and heart failure, with elevated rates specific to FASD females. In individuals with FASDs, CHD diagnosis was associated with an increased incidence of conduction defects, arrhythmias, heart remodeling, heart failure, and systolic and diastolic dysfunction. Increased CVD burden in FASD adults remained significant after adjustment for BMI, composite cardiometabolic abnormalities, and hyperlipidemia.

**Conclusions:** Adults with FASDs exhibit an increased burden of CVD not fully explained by conventional cardiometabolic risk factors. These findings support enhanced cardiovascular screening in individuals with FASDs.

## Introduction

Prenatal alcohol exposure (PAE) affects over 10% of newborns globally and leads to the development of fetal alcohol spectrum disorders (FASDs) (1). Approximately 2 – 5% of U.S. school age children have FASDs, which manifest in features such as cognitive and behavioral deficits, craniofacial anomalies, and organ malformations (2–4). While not often appreciated as an important target of PAE, the heart is particularly vulnerable to damage, especially during the first trimester when critical heart structures form. Despite this, comprehensive studies on the burden of congenital and age-related cardiovascular disease (CVD) in FASD patient populations are lacking. This represents a critical knowledge gap because unrecognized cardiovascular risk in FASD patients may delay preventive care and exacerbate long-term morbidity and mortality.

Over the last 50 years, multiple studies have reported a high prevalence of congenital heart defects (CHDs) in individuals with FASDs, including atrial septal defects (ASD), ventricular septal defects (VSD), and conotruncal malformations (5–7). However, estimates of CHD prevalence in FASD patient populations are inconsistent, likely due to historically small sample sizes and inclusion bias. Whereas some studies indicate that up to 70% of patients with fetal alcohol syndrome (FAS) and over 30% of patients with any FASD have a CHD (5–7), other studies have found minimal to no association between PAE and congenital cardiac anomalies (8–14). A recent large-scale evaluation of Medicaid (n = 8,732,345) and Commercial health insurance data (n = 10,567,765) from 2016-2022 revealed that CHDs are more common amongst children with FASDs than those without (5.2% vs. 1.0% (Medicaid); 3% vs. 0.5% (Commercial)) (15). Collectively, these studies indicate a need to clarify the extent to which CHDs are comorbid with FASDs, especially because many CHDs might not be adequately identified during routine infant screening and could require healthcare providers to prioritize cardiac assessments in individuals with suspected PAE.

Research on age-related CVD in individuals with FASDs is limited, in part due to the relative newness of FASDs as a clinical umbrella diagnosis. Despite longstanding historical observations of alcohol-related developmental effects, FAS was not formally recognized until 1973 (6), reducing the likelihood that current adults would have received a diagnosis and subsequently participated in age-related FASD assessments. In animal model research, embryonic alcohol exposure is associated with increased rates of cardiomyopathy and diastolic dysfunction, with age-related disease more readily assessed than in humans (16). In human studies, children and adolescents with FASDs have been found to have a higher incidence of hypertension (17). A self-report-based survey of adults with FASDs also indicated a higher incidence of cardiomyopathy than the general population (18). “Cardiovascular problems” were also identified as comorbid with FASDs in a hospital discharge database (19); however, specific cardiovascular conditions were not delineated. Isolated reports of conduction abnormalities in the absence of structural defects or channelopathies further support the hypothesis that PAE confers long-term vulnerability to diverse CVD phenotypes (20).

Clinical data registries, which contain detailed medical histories, can help identify adults with FASDs for retrospective studies that would otherwise be difficult to conduct prospectively. Using a research patient data registry (RPDR) from a large academic hospital, we previously identified >200 adults with FASDs and conducted a retrospective cross-sectional study demonstrating a higher prevalence of metabolic abnormalities, including type 2 diabetes, low HDL, high triglycerides, and female-specific overweight and obesity relative to matched controls (21). Though the higher rates of adverse cardiometabolic health conditions suggest potential increased risk for CVD, our prior study was not designed to assess longitudinal CVD outcomes. Leveraging our previously identified patient cohort of adults with FASDs with additional longitudinal follow-up, this study examines the prevalence of CHDs, age-related CVD incidence, and the associations between CVD and cardiometabolic outcomes. Given documentation of sex-specific phenotypes in our prior study, we assessed sex-by-diagnosis interactions to determine whether males and females are equally impacted by CVD.

## Methods

### Study Approval

The Mass General Brigham institutional review board approved the current study (2017P000752), and informed consent was waived.

### Identification of Study Cohort

Males and females with FASDs > 18 years old at the time of data review were identified (n = 208) using the Research Patient Data Registry (RDPR), an electronic health record (EHR) database comprising detailed health records for all patients seen at one of nine hospitals in the Mass General Brigham multi-institutional healthcare system (Fig. 1). FASD-suspected patients were first identified through a medical record search query for fetal alcohol or prenatal alcohol exposure, FASD, and FASD sub diagnoses. Manual review of the Electronic Health Records (EHR) was undertaken for all participants to confirm the presence of a FASD diagnosis and to collect available demographic data. Only individuals with a documented diagnosis of fetal alcohol syndrome (FAS), fetal alcohol effects (FAE), partial fetal alcohol syndrome (pFAS), alcohol-related birth defect (ARBD), or alcohol related neurodevelopmental disorder (ARND) were included in the FASD cohort. FASD cases were then matched on age and sex in a 1:3 ratio to controls without FASD, all of whom were also > 18 years old (n = 824). Approximately 25% of controls were identified from the RPDR, whereas the remaining 75% (n = 624) were derived from the Community Care Cohort Project (C3PO), an EHR-based cohort of 520,868 individuals receiving longitudinal primary care between 2001 and 2018 (22). By selecting individuals on the basis of routine primary care rather than specific data availability, the C3PO design has been previously shown to improve longitudinal ascertainment and reduce missingness and bias compared to standard approaches to EHR ascertainment (22). Manual review of the EHR was conducted for each control patient to confirm the absence of a FASD diagnosis or documented prenatal alcohol exposure. All variables used for primary analyses were also obtained via standardized manual EHR abstraction for FASD and control patients. A systematic search method was employed for manual data abstraction, which included review of the problem list, labs, clinic notes, specialty tabs (Cardiology and Imaging), and linked records from non-MGB institutions where available. The search function was used to further confirm presence or absence of an outcome, and to identify the date the outcome was first mentioned in the EHR. The last available follow-up date, corresponding to the last clinic visit date considered in the file for each individual, was defined as the cutoff date for abstraction of clinical information from the EHR. All diagnoses, laboratory values, imaging results, and clinical assessments occurring at or prior to the last available follow-up date were eligible for inclusion. Age was defined as of the last available follow-up date. A total of 7 control patients were excluded due to incomplete records, end-stage organ failure with markedly elevated baseline cardiovascular risk, or EHR documentation suggestive of prenatal alcohol exposure. Except for quantitative data, absence of a documented diagnosis or relevant imaging finding in the EHR was interpreted as absence of the corresponding condition. No imputation of missing values was performed. Male and female designation was defined by assigned sex at birth.

**Figure 1.**
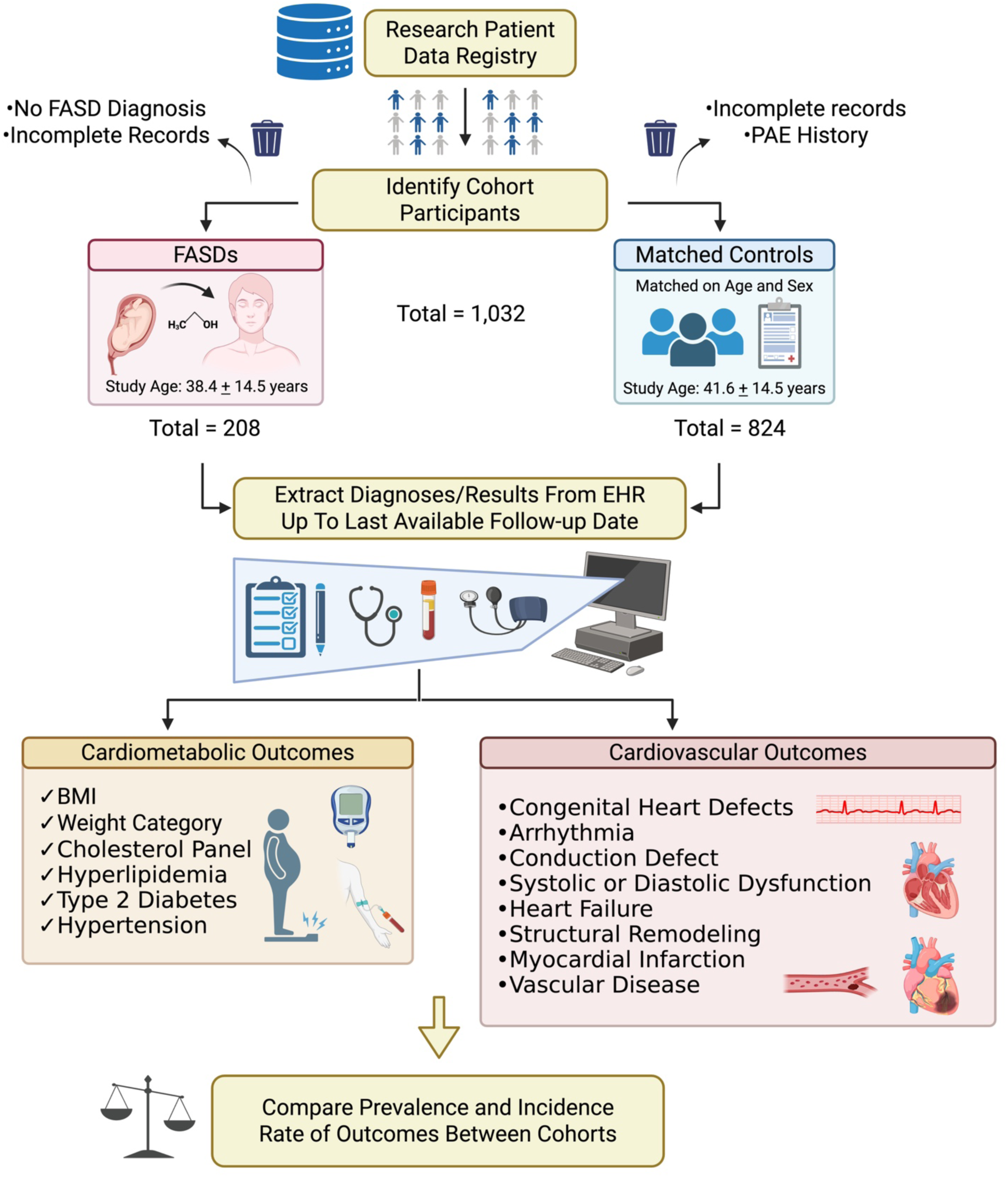
Overview of Cohort Study. Graphic overview of the retrospective clinic cohort study. FASD and matched control patients (n = 1,032) were identified through a large academic research data patient registry (RPDR). Cardiometabolic and cardiovascular outcomes were extracted from the EHR up to the last available follow-up date, which corresponds to the date of the last medical records considered in the EHR. Date of diagnosis was recorded for cardiovascular conditions, when available. Prevalence and incidence rates were compared between control and FASD cohorts. Created in BioRender. Weeks, O. (2026) https://BioRender.com/le8ap1l.

### Cardiometabolic Outcomes

The main cardiometabolic outcomes were body mass index (BMI), cholesterol level, hyperlipidemia, type 2 diabetes mellitus, and hypertension. Outcome data collected represent the most recent available data in the EHR at or prior to the last available follow-up date unless otherwise indicated. Quantitative BMI was defined as weight (kg)/height (m)^2^ and categorized as: BMI <18.5 = underweight; 18.5 – 24.9 = normal; 25.0 – 29.9 = overweight; >30.0 = obese. Blood levels of HDL cholesterol (mg/dL) and triglycerides (mg/dL) obtained as part of regular medical care were collected irrespective of antihyperlipidemic medication status. HDL cholesterol < 40 mg/dL, considered abnormally low, was a binary study outcome. Hyperlipidemia was defined as having a formal diagnosis of hyperlipidemia by a clinician, or quantitative evidence of elevated lipid panel results at any date prior to the last available follow-up date (total cholesterol ≥ 200 mg/dL or triglycerides >150 mg/dL or LDL > 130 mg/dL). Type 2 diabetes mellitus (T2DM) was defined as having a formal diagnosis made by a physician, quantitative documentation of ≥ 2 hemoglobin A1C measurements ≥ 6.5%, or ≥ 2 elevated fasting plasma glucose levels (reference range determined by reporting laboratory) in the absence of a type 1 diabetes mellitus diagnosis. Hypertension was defined as having a formal diagnosis of hypertension by a physician at or before the last available follow-up date, or documentation of blood pressure readings with both a systolic pressure >130 and a diastolic pressure of > 80 mmHg on two separate clinic visits. The presence of multiple cardiometabolic abnormalities was defined as the co-occurrence of overweight/obesity, low HDL cholesterol (< 40 mg/dL), and T2DM.

### Cardiovascular Outcomes

The main cardiovascular outcomes were congenital heart defect (CHD), heart murmur, arrhythmia, conduction defect, structural heart remodeling (wall thickening or chamber dilation), systolic or diastolic dysfunction, systolic or diastolic heart failure, myocardial infarction (MI), stroke/cerebral vascular accident, and thrombosis or embolism. Outcome data were manually collected from the EHR and represent all diagnoses and exams present at or prior to the last available follow-up date. CHD is defined as having a diagnosis of a congenital cardiac anomaly in clinical notes or an echocardiography exam report indicating a CHD finding. Minor defects, including congenital valve anomalies and patent foramen ovale (PFO), were included among the CHD endpoints, given variability in clinical documentation of small or previously repaired septal defects. Heart murmur was defined as having a physician note of a heart murmur in the EHR. Arrhythmia was defined as having a formal diagnosis of atrial or ventricular arrhythmia (atrial fibrillation, atrial flutter, supraventricular tachycardia, ventricular tachycardia, ventricular fibrillation) in the EHR or an EKG report or ambulatory monitor indicating an arrhythmia. Conduction defects (bundle branch block, fascicular block, heart block, and Braguda syndrome), which were categorized separately from arrhythmias, were defined as having a conduction defect diagnosis made by a physician or an EKG result indicating a conduction defect finding not attributable to acute illness. Wolff-Parkinson-White syndrome was classified as an arrhythmia, reflecting the clinical manifestation of pre-excitation-mediated tachyarrhythmia. Structural heart remodeling was defined as having an indication of (left or right) atrial enlargement or ventricle dilation or hypertrophy on an echocardiography report. Systolic heart failure was defined as having documented systolic heart failure or a confirmation of a left ventricular ejection fraction of ≤40%. Systolic dysfunction was defined as having documentation of systolic dysfunction in the EHR, reduced left ventricular ejection fraction (≤50%), or abnormal ventricular wall movement documented in an echocardiography report. Diastolic heart failure was defined as having a diagnosis of diastolic heart failure or heart failure with preserved ejection fraction (HFpEF) in the EHR. Diastolic dysfunction was defined as having a diagnosis of diastolic heart failure, HFpEF, or notes indicating diastolic dysfunction in an echocardiography report. To quantify left ventricular systolic function, we collected data on left ventricular ejection fraction (EF%), left ventricular end-diastolic internal dimension (LVIDd), and left ventricular end-systolic internal dimension (LVIDs). Quantitative values for intraventricular septum thickness (IVS) and posterior wall thickness (PWT) were also collected from the echocardiography report. When multiple echocardiography reports were available, quantitative data were retrieved from the echo closest to, but before, the last available follow-up date. For quantitative echocardiographic parameters reported as ranges (e.g. EF%, 60 – 65%), the midpoint of the range was used for analysis. MI, stroke/cerebral vascular accident, and thromboembolic events were defined as having a diagnosis of the condition in the EHR. Imaging studies were often available to corroborate the finding but were not required. Thromboembolic events included pulmonary embolism, deep vein thrombosis, and related thrombophlebitis.

### Statistics

Descriptive statistics included mean±SD, median with interquartile range or range, and frequency with percentage. Crude comparisons were performed with Student’s t-test for continuous outcomes and with a Fisher exact test for categorical outcomes. Analysis of covariance was performed to compare BMI and metabolic parameters with age at last available follow-up as the covariate, with age-adjusted mean ± standard error (SE) reported. This approach was also used to compare the prevalence of cardiovascular outcomes. For the specific outcomes of hypertension, stroke, and thromboembolism, the age covariate was the documented age at diagnosis and age at last available follow-up for those without the condition. Poisson regression was also performed to estimate cardiovascular condition incidence rates of the FASD and control groups, accounting for follow-up time. Incidence rates were calculated as the number of individuals with a documented condition divided by the sum of age-based person-time, where person-time was defined as age at condition onset. Participants without a documented diagnosis of a given condition were censored at their age at the last available follow-up date. Given documented sex differences in FASDs, tests of interaction (sex by cohort [FASD vs. control]) were performed to assess whether the FASD vs. control difference in mean cardiometabolic parameters and in incidence of conditions varied by sex. Unless otherwise noted, reported group comparisons reflect age-adjusted estimates. A p-value of 0.05 or smaller was considered statistically significant. Analyses were performed using SAS version 9.4 (SAS Institute, Inc., Cary, NC) and R 4.4.2 (23).

### Data Availability

Data will be shared via Data Use Agreements, subject to institutional policy and IRB approval.

## Results

### Study Participants

At the last available follow-up date, FASD participants were age 18.01 to 80.3 years (mean [SD] age, 38.4 [14.5] years; 36% aged < 30 years, 41% aged 30 to <50 years, 23% aged ≥ 50 years; 50% male and 50% female; Table 1, Fig. S1A-C). Control participants were aged 18.1 to 89.6 years (mean [SD] age, 41.6 [14.5] years; 22% aged < 30 years, 50% aged 30 to <50 years, 28% aged ≥ 50 years; 49% male and 51% female). Despite age matching, controls were slightly older than the FASD cohort at the last available follow-up date (p<0.001). Two-thirds of both groups had non-Hispanic white racioethnicity (66% for FASD and 67% for control, p=0.85; Table 1, Fig. S1D – G). The distributions of racial categories differed (p<0.001) between FASD and control cohorts. Of note, FASD participants were more likely to be Black/African American (15% vs. 10%) and less likely to be Asian (0% vs 4%).

**Table 1.** Demographic Characteristics.

|  | Combined |  |  | Males |  |  | Females |  |  |
| --- | --- | --- | --- | --- | --- | --- | --- | --- | --- |
| Personal | FASD | Control | P value | FASD | Control | P value | FASD | Control | P value |
| N | 208 | 824 |  | 103 | 406 |  | 105 | 418 |  |
| Age at last available follow-up, yrs | 38.4±14.5 | 41.6±14.5 | **0.004** | 38.0±14.7 | 41.6±15.1 | **0.03** | 38.7±14.3 | 41.7±13.9 | 0.06 |
| Age group, yrs |  |  | **<0.001** |  |  | **0.03** |  |  | **0.01** |
| < 30 | 75 (36.1%) | 185 (22.4%) |  | 36 (35.0%) | 90 (22.2%) |  | 39 (37.1%) | 95 (22.7%) |  |
| 30 to <50 | 86 (41.4%) | 410 (49.8%) |  | 45 (43.7%) | 208 (51.2%) |  | 41 (39.1%) | 202 (48.3%) |  |
| ≥ 50 | 47 (22.6%) | 229 (27.8%) |  | 22 (21.4%) | 108 (26.6%) |  | 25 (23.8%) | 121 (29.0%) |  |
| White, non-Hispanic<sup>*</sup> | 95 (66.0%) | 441 (67.1%) | 0.85 | 43 (59.7%) | 204 (63.8%) | 0.59 | 52 (72.2%) | 237 (70.3%) | 0.89 |
| Race |  |  | **<0.001** |  |  | **0.009** |  |  | 0.19 |
| N | 186 | 596 |  | 97 | 357 |  | 89 | 239 |  |
| White | 150 (77.3%) | 578 (77.5%) |  | 71 (73.2%) | 269 (75.4%) |  | 79 (81.4%) | 309 (79.4%) |  |
| Black/African American | 29 (15.0%) | 72 (9.7%) |  | 17 (17.5%) | 33 (9.2%) |  | 12 (12.4%) | 39 (10.0%) |  |
| Asian | 0 (0%) | 29 (3.9%) |  | 0 (0%) | 13 (3.6%) |  | 0 (0%) | 16 (4.1%) |  |
| Amer Indian/Alaskan Native | 1 (0.5%) | 0 (0%) |  | 1 (1.0%) | 0 (0%) |  | 0 (0%) | 0 (0%) |  |
| More than one race | 3 (1.6%) | 5 (0.7%) |  | 2 (2.1%) | 3 (0.8%) |  | 1 (1.0%) | 2 (0.5%) |  |
| Other | 11 (5.7%) | 62 (8.3%) |  | 6 (6.2%) | 39 (10.9%) |  | 5 (5.2%) | 23 (5.9%) |  |
| Hispanic ethnicity<sup>†</sup> | 15 (11.1%) | 91 (14.2%) | 0.41 | 10 (15.4%) | 55 (17.7%) | 0.72 | 5 (7.1%) | 36 (10.8%) | 0.51 |

<sup>\*</sup>Reduced sample size due to missing data. n=72 (FASD male); 320 (CTRL male); 72 (FASD female); 337 (FASD female).
<sup>†</sup>Reduced sample size due to missing data. n=65 (FASD male); 311 (CTRL male); 70 (FASD female); 332 (FASD female).
FASD=Fetal alcohol spectrum disorder.

### Cardiometabolic Health

We observed an interaction between sex and BMI (p <0.001) and sex and weight category (p <0.001; Table 2). FASD males had no difference in mean [SE] BMI relative to controls (27.1±0.8 vs. 28.6±0.5; p = 0.15; Table 2, Fig. S2A). In contrast, FASD females had higher mean BMI than controls (31.0±0.8 vs. 27.2±0.4; p = <0.001; Table 2, Fig. S2B). FASD males had a lower prevalence of overweight and obesity (p = 0.007) whereas FASD females had a higher prevalence of overweight and obesity than female controls (p = 0.002; Table 2).

**Table 2.**
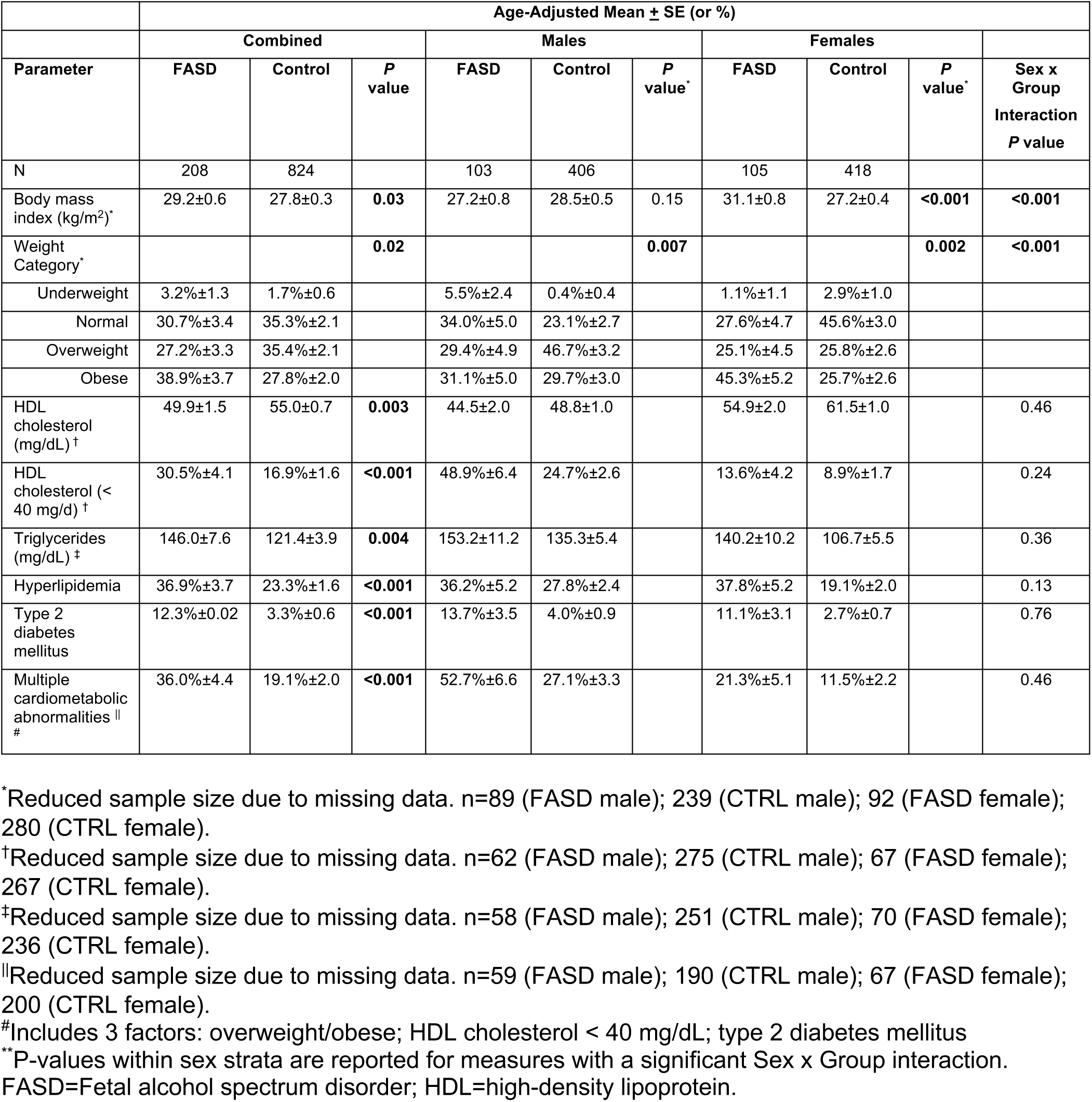
Cardiometabolic Parameters at Age of Last Follow-Up.

We next examined dyslipidemia metrics, including HDL cholesterol (mg/dL), triglycerides (mg/dL), and hyperlipidemia diagnosis. Adults with FASDs had lower HDL cholesterol levels than controls (49.9 ±1.5 vs. 55.0±0.7; p = 0.003) and a higher prevalence of low HDL (<40 mg/dL; 31<u>%</u> vs. 17%; p <0.001; Table 2, S1, Fig. S2C – D). Triglyceride levels were higher (146.0±7.6 vs. 121.4±3.9; p = 0.004), and hyperlipidemia (37% vs. 23%; p <0.001), T2DM (12% vs. 3%; p < 0.001), and clustering of multiple cardiometabolic abnormalities (36% vs. 19%; p < 0.001) were also more common in the FASD cohort relative to controls (Table 2, S1, Fig. S2E–F).

### Congenital Heart Defects

CHD diagnosis was more prevalent in individuals with FASD than controls (6% vs. 1%; p < 0.001, Table 3, Fig. 2A), with similar increases observed in FASD males and females (Males: 7% vs. 1%, p = 0.002; Females: 6% vs. 1%, p = 0.02; Table 3). CHD subtypes observed in the FASD cohort included septal defects, valve defects, patent ductus arteriosus, and conotruncal malformations. The prevalence of any one subtype of CHD was low; however, atrial septal defect (ASD; p = 0.06), ventricular septal defect (VSD; p = 0.11), and Tetralogy of Fallot (TOF) (p = 0.04) were more common in the FASD cohort (Table 3). Heart murmur prevalence was also higher in the FASD cohort but not statistically significant (5% vs. 3%; p = 0.07).

**Figure 2.**
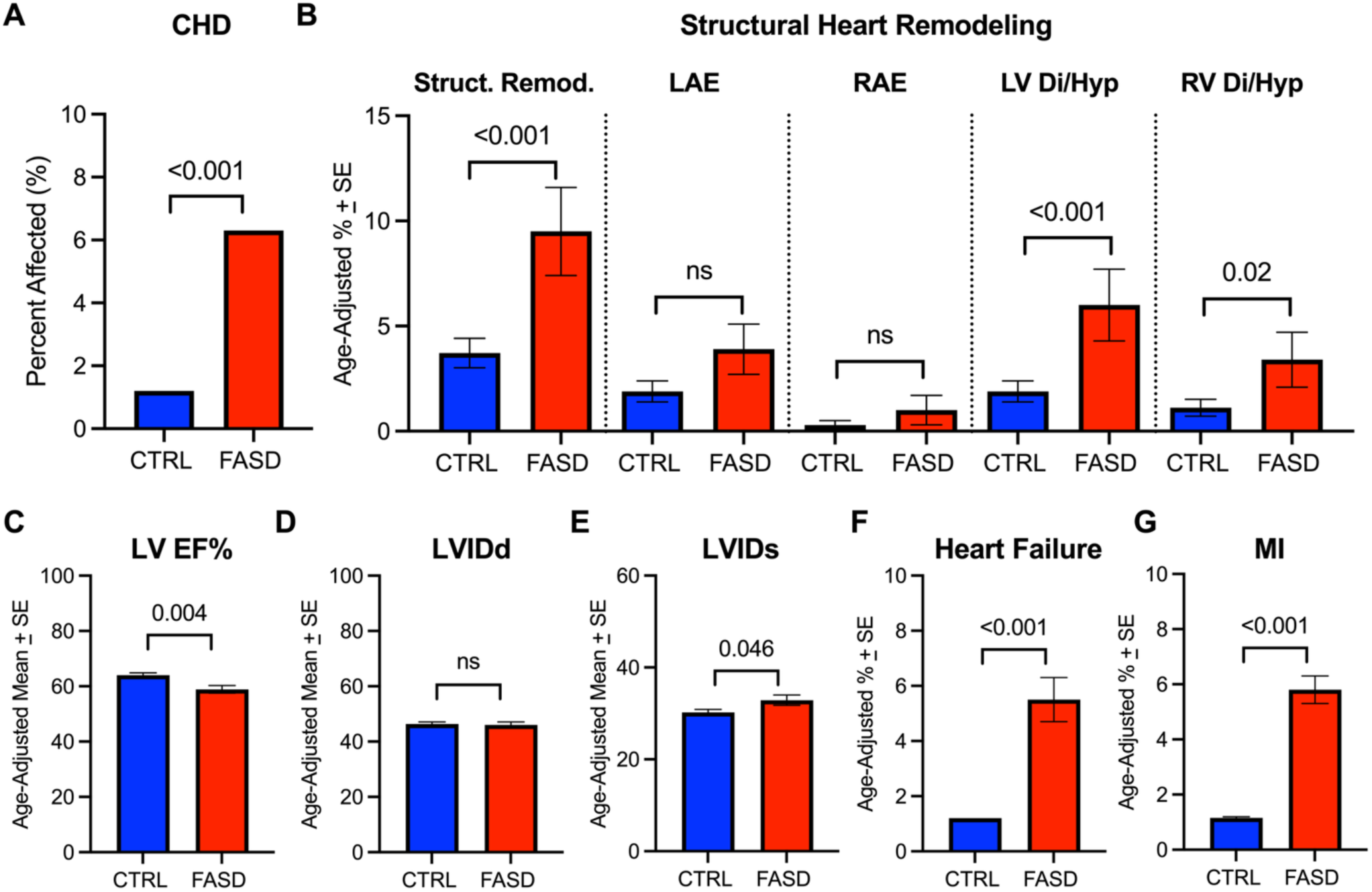
Cardiovascular disease prevalence in adults with FASDs. **A.** Prevalence of congenital heart defects (CHDs) is increased in FASDs. **B.** Prevalence of structural remodeling (struct. remod.), left atrial enlargement (LAE), right atrial enlargement (RAE), left ventricular dilation or hypertrophy (LV Di/Hyp), and right ventricular dilation or hypertrophy (RV Di/Hyp). Ventricular remodeling is more common in FASDs. **C.** Mean left ventricular ejection fraction (LV EF%) is reduced in FASDs, indicating reduced systolic function. **D.** Left ventricular end-diastolic internal dimension (LVIDd) is unaffected in FASDs. **E.** Left ventricular end-systolic internal dimension. (LVIDs) is increased in FASDs, indicating reduced systolic function. **F – G..** Prevalence of heart failure (F) and myocardial infarction (MI, G) is increased in FASDs. Data is from the combined cohort of males and females. Error bars represent standard error. See Table S3 and Table S4 for corresponding age-adjusted data. CTRL=control; FASD=fetal alcohol spectrum disorder.

**Table 3.** Congenital Heart Defects.

|  | Combined |  |  | Males |  | Females |  |  |
| --- | --- | --- | --- | --- | --- | --- | --- | --- |
| Defect | FASD | Control | *P* value | FASD | Control | FASD | Control | Sex x Group<br>Interaction<br>*P* value |
| N | 208 | 824 |  | 103 | 406 | 105 | 418 |  |
| **Any CHD diagnosis** | 13 (6.3%) | 10 (1.2%) | **<0.001** | 7 (6.8%) | 4 (1.0%) | 6 (5.7%) | 6 (1.4%) | 0.51 |
| Patent Foramen Ovale (PFO) | 3 (1.4%) | 4 (0.5%) | 0.15 | 2 (1.9%) | 1 (0.3%) | 1 (1.0%) | 3 (0.7%) |  |
| Atrial septal defect (ASD) | 4 (1.9%) | 4 (0.5%) | 0.06 | 1 (1.0%) | 1 (0.3%) | 3 (2.9%) | 3 (0.7%) |  |
| Bicuspid aortic valve | 1 (0.5%) | 4 (0.5%) | 1.00 | 1 (1.0%) | 1 (0.3%) | 0 | 3 (0.7%) |  |
| Ventricular septal defect (VSD) | 2 (1.0%) | 1 (0.1%) | 0.11 | 1 (1.0%) | 1 (0.3%) | 1 (1.0%) | 0 |  |
| Coarctation of the aorta | 0 (0%) | 2 (0.2%) | 1.00 | 0 | 2 (0.5%) | 0 | 0 |  |
| Tetralogy of Fallot (TOF) | 2 (1.0%) | 0 (0%) | **0.04** | 1 (1.0%) | 0 | 1 (1.0%) | 0 |  |
| Patent ductus arteriosus (PDA) | 1 (0.5%) | 0 (0%) | 0.20 | 0 | 0 | 1 (1.0%) | 0 |  |
| Transposition of the great arteries (TGA) | 1 (0.5%) | 0 (0%) | 0.20 | 1 (1.0%) | 0 | 0 | 0 |  |
| Aortic stenosis | 0 (0%) | 2 (0.2%) | 1.00 | 0 | 1 (0.3%) | 0 | 1 (0.2%) |  |
| **Other diagnosis** |  |  |  |  |  |  |  |  |
| Heart Murmur | 11 (5.3%) | 22 (2.7%) | 0.07 | 4 (3.9%) | 9 (2.2%) | 7 (6.7%) | 13 (3.1%) | 0.78 |

FASD=Fetal alcohol spectrum disorder; CHD=congenital heart defect.

### Hypertension

A sex-by-diagnosis interaction was observed for hypertension (p = 0.01; Table 4). FASD females, but not males, had a higher incidence rate of hypertension relative to controls (Females: 0.78±0.14 vs. 0.41±0.05, p = 0.004; Males: 0.58±0.12 vs 0.69±0.06, p = 0.47; Table 4). Age-adjusted prevalence using logistic regression identified a significant increase in hypertension in the sex-combined FASD cohort relative to controls (Table S2 – S3).

**Table 4.** Age-Adjusted Incidence Rates per 100 patient-years±SE of Hypertension and Adult Cardiovascular Diseases by Sex.

|  | Age-Adjusted Incidence Rate per 100 patient-years±SE |  |  |  |  |  |  |  |  |  |  |  |  |
| --- | --- | --- | --- | --- | --- | --- | --- | --- | --- | --- | --- | --- | --- |
|  | COMBINED |  |  |  | MALES |  |  |  | FEMALES |  |  |  |  |
| Condition | FASD | Control | Rate Ratio<br>(95% CI) | P value | FASD | Control | Rate Ratio<br>(95% CI) | P value<sup>‡</sup> | FASD | Control | Rate Ratio<br>(95% CI) | P value<sup>‡</sup> | Interact.<br>P value |
| N | 208 | 824 |  |  | 103 | 406 |  |  | 105 | 418 |  |  |  |
| Hypertension | 0.68±0.09 | 0.55±0.04 | 1.24<br>(0.91, 1.69) | 0.17 | 0.58±0.12 | 0.69±0.06 | 0.85<br>(0.53, 1.33) | 0.47 | 0.78±0.14 | 0.41±0.05 | 1.88<br>(1.23, 2.88) | **0.004** | **0.01** |
| Any cardiac condition *<br>(excluding HTN) | 0.63±0.09 | 0.27±0.03 | 2.36<br>(1.67, 3.34) | **<0.001** | 0.61±0.13 | 0.32±0.04 | 1.92<br>(1.19, 3.10) | **0.008** | 0.64±0.13 | 0.21±0.03 | 3.01<br>(1.82, 4.97) | **<0.001** | 0.20 |
| Conduction defect * | 0.19±0.05 | 0.04±0.01 | 4.30<br>(2.10, 8.79) | **<0.001** | 0.20±0.07 | 0.04±0.02 | 4.93<br>(1.79, 13.61) | **0.002** | 0.17±0.07 | 0.05±0.02 | 3.75<br>(1.36, 10.33) | **0.01** | 0.71 |
| Cardiac arrhythmia * | 0.21±0.05 | 0.08±0.02 | 2.52<br>(1.38, 4.59) | **0.003** | 0.18±0.07 | 0.13±0.03 | 1.37<br>(0.59, 3.22) | 0.46 | 0.25±0.08 | 0.04±0.02 | 6.11<br>(2.33, 16.06) | **<0.001** | **0.02** |
| Atrial fibrillation | 0.10±0.03 | 0.05±0.01 | 2.01<br>(0.87, 4.68) | 0.10 | 0.13±0.06 | 0.09±0.02 | 1.43<br>(0.52, 3.93) | 0.49 | 0.07±0.04 | 0.01±0.01 | 6.44<br>(1.08, 38.52) | **0.04** | 0.15 |
| Structural heart remodeling* | 0.28±0.06 | 0.13±0.02 | 2.20<br>(1.32, 3.68) | **0.003** | 0.33±0.09 | 0.19±0.03 | 1.75<br>(0.92, 3.34) | 0.09 | 0.22±0.07 | 0.06±0.02 | 3.50<br>(1.45, 8.45) | **0.005** | 0.21 |
| Left atrial enlargement * | 0.14±0.04 | 0.08±0.02 | 1.69<br>(0.84, 3.39) | 0.14 | 0.15±0.06 | 0.12±0.03 | 1.30<br>(0.52, 3.23) |  | 0.12±0.06 | 0.05±0.02 | 2.68<br>(0.88, 8.18) |  | 0.32 |
| Right atrial enlargement * | 0.04±0.02 | 0.01±0.01 | 3.22<br>(0.72, 14.40) | 0.13 | 0.05±0.04 | 0.02±0.01 | 2.88<br>(0.48, 17.23) |  | 0.02±0.02 | 0.01±0.01 | 4.28<br>(0.27, 68.43) |  | 0.81 |
| LV dilation/hypertrophy * | 0.20±0.05 | 0.08±0.02 | 2.46<br>(1.33, 4.54) | **0.004** | 0.26±0.08 | 0.13±0.03 | 1.96<br>(0.93, 4.14) |  | 0.15±0.06 | 0.03±0.01 | 4.28<br>(1.38, 13.27) |  | 0.26 |
| RV dilation/hypertrophy * | 0.09±0.03 | 0.03±0.01 | 3.34<br>(1.24, 8.97) | **0.02** | 0.13±0.06 | 0.05±0.02 | 2.69<br>(0.88, 8.24) |  | 0.05±0.03 | 0.01±0.01 | 8.56<br>(0.78, 94.41) |  | 0.39 |
| Heart Failure (HF)* | 0.15±0.04 | 0.04±0.01 | 3.44<br>(1.61, 7.35) | **0.001** | 0.13±0.05 | 0.07±0.02 | 1.80 | 0.27 | 0.17±0.07 | 0.02±0.01 | 9.99 | **<0.001** | **0.05** |
|  | COMBINED |  |  |  | MALES |  |  |  | FEMALES |  |  |  |  |
| Condition | FASD | Control | Rate Ratio<br>(95% CI) | P value | FASD | Control | Rate Ratio<br>(95% CI) | P value<sup>†</sup> | FASD | Control | Rate Ratio<br>(95% CI) | P value<sup>†</sup> | Interact.<br>P value |
|  |  |  |  |  |  |  | (0.63,<br>5.11) |  |  |  | (2.58,<br>38.62) |  |  |
| Systolic HF * | 0.11±0.04 | 0.03±0.10 | 3.52<br>(1.46, 8.49) | **0.005** | 0.15±0.06 | 0.02±0.04 | 2.35<br>(0.87,<br>6.37) | 0.09 | 0.07±0.04 | 0 | - | - | - |
| Diastolic HF* | 0.09±0.03 | 0.02±0.01 | 3.76<br>(1.36,<br>10.37) | **0.01** | 0.04±0.01 | 0.02±0.01 | 2.16<br>(0.40,<br>11.79) |  | 0.13±0.06 | 0.02±0.01 | 5.35<br>(1.44,<br>19.93) |  | 0.41 |
| Systolic or Diastolic<br>Dysfunction* | 0.16±0.05 | 0.06±0.01 | 2.94<br>(1.45, 5.96) | **0.003** | 0.15±0.06 | 0.09±0.02 | 1.73<br>(0.67,<br>4.45) |  | 0.17±0.07 | 0.02±0.02 | 7.49<br>(2.19,<br>25.59) |  | 0.06 |
| Myocardial infarction | 0.15±0.04 | 0.04±0.01 | 4.33<br>(1.95, 9.65) | **<0.001** | 0.18±0.07 | 0.07±0.02 | 2.54<br>(1.00,<br>6.45) |  | 0.12±0.06 | 0 | - |  | - |
| Stroke/Cerebral Vascular<br>Accident | 0.08±0.03 | 0.02±0.01 | 4.36<br>(1.41,<br>13.51) | **0.01** | 0.05±0.04 | 0.02±0.01 | 2.16<br>(0.49,<br>11.80) |  | 0.10±0.05 | 0.01±0.01 | 8.79<br>(1.61,<br>47.97) |  | 0.25 |
| Ischemic *<sup>†</sup> | 0.04±0.02 | 0.02±0.01 | 2.19<br>(0.55, 8.76) | 0.27 | 0.03±0.004 | 0.02±0.01 | 1.08<br>(0.12,<br>9.66) |  | 0.05±0.04 | 0.01±0.01 | 4.45<br>(0.63,<br>31.56) |  | 0.35 |
| Hemorrhagic * | 0.01±0.01 | 0 | Non-est. | - | 0.03±0.03 | 0 | - |  | - | 0 | 0 |  | 1.00 |
| Thrombosis or embolism | 0.11±0.04 | 0.03±0.01 | 4.33<br>(1.72,<br>10.90) | **0.002** | 0.05±0.04 | 0.03±0.01 | 1.73<br>(0.34,<br>8.93) |  | 0.17±0.07 | 0.02±0.01 | 7.56<br>(2.21,<br>25.82) |  | 0.16 |

\* The incidence rates for these conditions may be underestimates. No age at onset was available for these conditions, so age at last available follow-up was used instead.
<sup>†</sup>Reduced sample size due to missing data on type of stroke. n=103 (FASD male); 406 (CTRL male); 101 (FASD female); 418 (FASD female).
<sup>‡</sup>**P-values** within sex strata are reported for measures with a significant Sex x Group interaction.
FASD=Fetal alcohol spectrum disorder; HTN=hypertension; LV=left ventricle; RV=right ventricle; HF=heart failure.

### Conduction Defect and Arrhythmia

The incidence rate of conduction defects was significantly enriched in the FASD cohort, with similar effects across sex (0.19±0.05 vs. 0.04±0.01 cases per 100 patient-years; rate ratio 4.30, 95% confidence interval 2.10 to 8.79; p < 0.001; Table 4). Conduction defects observed in the FASD cohort included intraventricular conduction defect (left and right bundle branch block and left anterior fascicular block) and first-degree atrioventricular block. A significant interaction between sex and arrhythmia was observed, with FASD females but not males having higher arrhythmia incidence than controls (Females: 0.25±0.08 vs. 0.04±0.02 cases per 100 patient-years, p < 0.001; Males: 0.18±0.07 vs. 0.13±0.03 cases per 100 patient-years), p = 0.46; Table 4). Arrhythmias in the FASD cohort included ventricular tachycardia, ventricular bigeminy, and atrial fibrillation. One FASD patient had documented Wolff-Parkinson-White Syndrome. The incidence of atrial fibrillation, the most common type of arrhythmia in adults, was higher in FASD than control cohorts, though results were not significant (0.10±0.03 vs. 0.05±0.01 cases per 100 patient-years; p = 0.10; Table 4). A sensitivity analysis using logistic regression–derived, age-adjusted prevalence estimates yielded consistent results, confirming the incidence rate analysis (Table S2–S3).

### Structural Heart Remodeling and Quantitative Echocardiography Parameters

FASD adults had a higher incidence rate of structural heart changes relative to controls (0.28±0.06 vs. 0.13±0.02 cases per 100 patient-years; p = 0.003; Table 4). Incidence rates of left and right atrial enlargement were similar between cohorts; however, left and right ventricular remodeling was more frequent in individuals with FASD than controls (Left: 0.20±0.05 vs. 0.08±0.02 cases per 100 patient-years; p = 0.004; Right: 0.09±0.03 vs. 0.03±0.01 cases per 100 patient-years; p = 0.02). No interaction between sex and structural heart remodeling was observed (p = 0.21). A sensitivity analysis of age-adjusted prevalence estimates confirmed increased structural heart remodeling in adults with FASDs (Fig. 2B, Table S2–S3).

Adults with FASD had lower mean (±SE) EF% than controls (58.9±1.4 vs. 64.0±0.9; p = 0.004; Fig. 2C, Table S4, Fig. S3A – C). Mean (±SE) LVIDs, but not LVIDd, was higher in the FASD cohort than controls, indicating reduced contractility during systole (32.9±1.1 vs. 30.2±0.7; p =0.046; Fig, 2D – E, Table S4, Fig. S3D – E). IVS and PWT were unaffected by FASD status (p > 0.05; Table S4, Fig. S3F – G). No sex-by-diagnosis interactions were observed for echocardiography parameters.

### Heart Failure and Myocardial Infarction

Heart failure incidence was higher in the FASD cohort than in controls (p = 0.001; Table 4), with a significant sex-by-diagnosis interaction (p = 0.05). FASD females, but not males, had a higher incidence of heart failure than controls (females: 0.17±0.07 vs. 0.02±0.01 cases per 100 patient-years; p < 0.001). Systolic and diastolic dysfunction (with or without heart failure) were also more frequent in the FASD cohort relative to controls (0.16±0.05 vs. 0.06±0.01 cases per 100 patient-years; p = 0.003; Table 4). Myocardial infarction (MI) similarly had a higher incidence rate in individuals with FASDs (0.15±0.04 vs. 0.04±0.01 cases per 100 patient-years; p < 0.001; Table 4). A sensitivity analysis of age-adjusted prevalence estimates confirmed increased heart failure and MI in adults with FASDs (Fig. 2F-G, Table S2–S3).

### Vascular Disease

Stroke incidence rate was higher in the FASD cohort than in controls (0.08±0.03 vs. 0.02±0.01 cases per 100 patient-years; p = 0.01; Table 4). Both ischemic and hemorrhagic strokes were observed in the FASD cohort, but most strokes were ischemic. Thromboembolic events were similarly increased in the FASD cohort relative to controls (0.11±0.04 vs. 0.03±0.01 cases per 100 patient-years; p = 0.002; Table 4). FASD females, but not males, had a significantly higher incidence rate of stroke (0.10±0.05 vs. 0.01±0.01 cases per 100 patient-years; p = 0.01) and thromboembolic events than controls (0.17±0.07 vs. 0.02±0.01 cases per 100 patient-years; p = 0.001); however, the sex-by-diagnosis interaction was not significant for stroke (p = 0.25) or for thromboembolic events (p = 0.16). A sensitivity analysis of age-adjusted prevalence estimates confirmed increased stroke and thromboembolism in adults with FASDs (Table S2–S3).

### Adult Cardiovascular Outcomes Stratified by CHD and Cardiometabolic Risk

The age-adjusted incidence rate of any cardiac condition (conduction defects, arrhythmias, structural heart remodeling, heart failure, systolic or diastolic dysfunction, MI, stroke, or thromboembolic events; excluding hypertension) in the FASD group was more than double that of the control group (0.63±0.09 vs. 0.27±0.03 cases per 100 patient-years; rate ratio 2.36, 95% confidence interval 1.67 to 3.34; p <0.001; Table 4). There was no sex-by-diagnosis interaction: the increased incidence of cardiac conditions with FASD was observed for both males and females (Female: 0.64±0.13 vs. 0.21±0.03 cases per 100 patient-years; Male: 0.61±0.0.13 vs. 0.32±0.04 cases per 100 patient-years).

We sought to determine whether CHD status or cardiometabolic abnormalities were associated with additional cardiovascular conditions within the FASD cohort, as identification of such correlates could inform risk stratification among individuals with FASD. FASD individuals with and without a CHD had a comparable mean age at last available follow-up (p=0.92; Table S5); therefore, unadjusted analyses are presented. In the FASD cohort, CHD diagnosis was associated with an increased prevalence of conduction defects (46% vs. 5%; p < 0.001), arrhythmias (46% vs. 6%; p < 0.001), structural heart remodeling (62% vs. 7%; p < 0.001), heart failure (31% vs. 4%; p = 0.004), and systolic or diastolic dysfunction (31% vs. 5%; p < 0.005; Table 5). In contrast, CHD status was not associated with hypertension, MI, stroke, or thromboembolic events (p > 0.05).

**Table 5.** Cardiovascular Diseases by CHD Status in the FASD Cohort.

| FASD Cohort | CHD |  |  |
| --- | --- | --- | --- |
| Condition | No | Yes | *P value* |
| N | 195 | 13 |  |
| Hypertension | 48 (24.6%) | 4 (30.8%) | 0.74 |
| Any cardiac condition (excluding HTN) | 37 (19.0%) | 13 (100%) | **<0.001** |
| Conduction defect | 9 (4.6%) | 6 (46.2%) | **<0.001** |
| Cardiac arrhythmia | 11 (5.6%) | 6 (46.2%) | **<0.001** |
| Atrial fibrillation | 5 (2.6%) | 3 (23.1%) | **0.009** |
| Structural heart remodeling | 14 (7.2%) | 8 (61.5%) | **<0.001** |
| Left atrial enlargement | 8 (4.1%) | 3 (23.1%) | **0.024** |
| Right atrial enlargement | 2 (1.0%) | 1 (7.7%) | 0.18 |
| LV dilation/hypertrophy | 10 (5.1%) | 6 (46.2%) | **<0.001** |
| RV dilation/hypertrophy | 2 (1.0%) | 5 (38.5%) | **<0.001** |
| Heart Failure (HF) | 8 (4.1%) | 4 (30.8%) | **0.004** |
| Systolic HF | 7 (3.6%) | 2 (15.4%) | 0.10 |
| Diastolic HF | 5 (2.6%) | 2 (15.4%) | 0.06 |
| Systolic or Diastolic Dysfunction | 9 (4.6%) | 4 (30.8%) | **0.005** |
| Myocardial infarction | 10 (5.1%) | 2 (15.4%) | 0.17 |
| Stroke/Cerebral Vascular Accident | 6 (3.1%) | 2 (15.4%) | 0.08 |
| Ischemic* | 2 (1.0%) | 1 (8.3%) | 0.17 |
| Hemorrhagic* | 1 (0.5%) | 0 (0%) | 1.00 |
| Thrombosis or embolism | 9 (4.6%) | 0 (0%) | 1.00 |

\*Reduced sample size due to missing data on type of stroke. n=192 (No CHD); 12 (Yes CHD).
FASD=Fetal alcohol spectrum disorder; CHD=congenital heart disease; HTN=hypertension; LV=left ventricular; RV=right ventricular; HF=heart failure.

Individuals with FASDs had higher odds of any cardiovascular condition (excluding hypertension) compared with controls (unadjusted OR: 2.55, 95% CI: 1.73–3.75; p<0.001; Table 6, S6). This association remained significant after adjustment for BMI, composite metabolic risk factors (overweight/obese, HDL cholesterol < 40 mg/dL, and type 2 diabetes mellitus), and hyperlipidemia, with adjusted odds ratios ranging from 2.17 to 2.43 (all p≤0.002; Table 6, S6). FASD was also associated with increased odds of structural heart remodeling (unadjusted OR: 2.15, 95% CI: 1.25–3.68; p=0.005; Table 6, S6). The association remained significant after adjustment for BMI (OR: 2.11, 95% CI: 1.25, 3.68; p=0.02) and hyperlipidemia (OR: 2.02, 95% CI: 1.17, 3.47; p=0.01) but was attenuated and no longer statistically significant after adjustment for multiple metabolic conditions (OR: 1.71, 95% CI: 0.86, 3.39; p=0.13; Table 6, S6).

**Table 6.** Covariate-Adjusted Models for the Presence of Any Cardiovascular Condition (Excluding Hypertension) or Heart Remodeling in FASD vs. Controls.

| **Any Cardiovascular Condition (Excluding Hypertension)** |  |  |
| --- | --- | --- |
| **Model** | **Odds Ratio (95% CI)** | **p-value** |
| FASD vs. Control (unadjusted) | 2.55 (1.73, 3.75) | <0.001 |
| FASD vs. Control | 2.29 (1.49, 3.52) | <0.001 |
| Body mass index (kg/m<sup>2</sup>) | 1.01 (0.99, 1.04) | 0.34 |
| FASD vs. Control | 2.17 (1.33, 3.54) | 0.002 |
| Multiple cardiometabolic abnormalities* | 1.72 (1.04, 2.85) | 0.03 |
| FASD vs. Control | 2.43 (1.65, 3.59) | <0.001 |
| Hyperlipidemia | 1.98 (1.36, 2.86) | <0.001 |
| **Structural Heart Remodeling** |  |  |
| **Model** | **Odds Ratio (95% CI)** | **p-value** |
| FASD vs. Control (unadjusted) | 2.15 (1.25, 3.68) | 0.005 |
| FASD vs. Control | 2.11 (1.14, 3.90) | 0.02 |
| Body mass index (kg/m<sup>2</sup>) | 1.04 (1.01, 1.08) | 0.008 |
| FASD vs. Control | 1.71 (0.86, 3.39) | 0.13 |
| Multiple cardiometabolic abnormalities* | 4.01 (2.07, 7.77) | <0.001 |
| FASD vs. Control | 2.02 (1.17, 3.47) | 0.01 |
| Hyperlipidemia | 2.18 (1.31, 3.63) | 0.003 |

\*Includes 3 factors: overweight/obese; HDL cholesterol<40; type 2 diabetes mellitus FASD=Fetal alcohol spectrum disorder

## Discussion

FASD patients who share their living experiences often express concern about the lack of information on long-term health trajectories, particularly with respect to cardiovascular outcomes. Moreover, they report cardiovascular symptoms that are unexplained and underrecognized within existing clinical frameworks. A primary goal of this study was to assess the burden of CHDs and CVD in a contemporary adult FASD patient population and to provide detailed information on cardiac chamber remodeling and quantitative measures of cardiac function, such as ejection fraction, which have been lacking from prior reports. We further sought to clarify whether CHD status or cardiometabolic abnormalities were associated with adverse cardiovascular outcomes as these phenotypes may aid in risk stratification.

We found that approximately 6 - 7% of adults with FASDs had a documented CHD, compared to 1% of controls. The prevalence of CHDs observed in our cohort is lower than earlier reports suggesting rates of 30 – 70% in FASD populations (5–7), but is more consistent with a recent analysis showing CHD prevalence of 5% among children with FASD insured by Medicaid and 3% among those with commercial insurance (15). Our findings confirm that PAE represents an underrecognized risk factor for CHD. In pregnant individuals with PAE, a fetal echocardiogram may be warranted to ensure timely CHD diagnosis. Furthermore, it may be valuable to screen individuals with a CHD for PAE and FASD, even when a genetic etiology for the CHD has been identified. Given that 2 - 5% of the U.S. population is estimated to have a FASD, and that up to 5 - 7% of individuals with FASD may have a CHD, PAE or gene-alcohol interactions could account for a meaningful fraction of CHDs (up to 10%).

Our findings further demonstrate individuals with FASDs had a higher risk of conduction defects and arrhythmias, structural heart remodeling, systolic and diastolic dysfunction, heart failure, MI, and vascular disease. Overall, FASD was associated with more than a two-fold increase in the odds of CVD (excluding hypertension), and this association persisted after adjustment for BMI, hyperlipidemia, and composite metabolic risk factors. These findings reveal that cardiometabolic abnormalities alone do not fully explain the elevated CVD risk in the adult FASD population. Instead, PAE may have direct and long-lasting impacts on the heart and vasculature, such as through epigenetic modifications and changes to cardiovascular development, that contribute to increased CVD risk. Comprehensive longitudinal cardiovascular screening is not currently recommended for individuals with PAE or FASDs; however, the burden of CVD in this FASD cohort suggests that current guidelines may warrant reevaluation. Routine exams, such as echocardiography and EKG, may help identify early problems with heart structure or function and increase the opportunity for more effective prevention. Additionally, prospective studies are needed to evaluate the effectiveness of early cardiovascular monitoring in individuals with FASDs, and to define how FASD screening can be optimally integrated into routine cardiovascular risk assessment.

Our findings prioritize recognition of FASDs as a potential key component of cardiovascular risk stratification. PAE screening and FASD diagnosis is currently limited and often hindered by stigma, patient heterogeneity, legal implications, and incomplete exposure histories. Estimates suggest that <1% of individuals a with FASD globally will receive a diagnosis (24). Given the impact of FASD on CHD and later CVD, incorporation of PAE and FASD screening into routine clinical care and EHR record documentation may improve longitudinal risk assessment and management.

## Strengths and Limitations

Several important strengths of this study merit recognition. To our knowledge, it is one of the first to systematically evaluate congenital and acquired CVDs in adults with FASDs. Leveraging a retrospective cohort design allowed examination of age-related cardiovascular outcomes across a wide adult age range (18 – 90 years), with a mean age of 38 in the FASD group and 42 years in controls. Another strength is the use of detailed EHRs through a large research data registry, enabling comprehensive validation of cardiovascular diagnosis through clinical documentation, electrocardiograms, and echocardiography reports. This level of phenotypic validation extends significantly beyond what is typically possible in studies that rely solely on administrative health data.

Several limitations warrant discussion. First, due to the retrospective nature of this study, identification of FASD and PAE was based on existing clinical documentation rather than prospective screening. Patients were not uniformly screened for PAE or FASD by the study team, and as a result, a small proportion of the control cohort may have undiagnosed FASDs, which would bias observed group differences toward the null. Moreover, cohort participants with a FASD diagnosis tended to have more severe forms of the condition (FAS); hence the effect sizes observed may be generalizable only to individuals with FAS or pFAS. While current diagnostic frameworks for FASD allow for patient stratification by sub-diagnosis (FAS, pFAS, ARND, ARBD), stratification was not feasible in this cohort due to variability in diagnostic tools and terms used in the EHR. Future studies would ideally have a strategy for screening control patients for PAE and should also examine the incidence of CVDs in patients with a broader range of FASDs.

Second, because of the retrospective design, age of disease onset could not be determined for all cardiovascular conditions, particularly among those who entered the EHR with pre-existing diagnoses. As a result, the incidence rates for several cardiovascular outcomes may be underestimates because, for these individuals, age at last available follow-up date was used instead of age of onset. Third, although sex-specific interactions were examined, the study was not powered primarily to detect interaction effects, and some clinically relevant sex differences may not have reached statistical significance. Fourth, our models may be affected by residual confounding, including potential effects of socioeconomic and behavioral factors, such as smoking and substance use, which were not controlled for in this study. These data may influence cardiovascular risk and should be incorporated into future studies when reliable data is available.

Lastly, not all participants underwent uniform cardiovascular testing. Those with echocardiography data may not be representative of the cohort, as testing was often limited to patients with more severe conditions. Even if the cohort with echocardiography data was representative, missing data for many patients resulted in limited statistical power. Prospective studies with systematic cardiovascular phenotyping across the lifespan will be critical to further define cardiac structure and function in adults with FASDs.

## Acknowledgements

Conceptualization: O.W., C.G.B., C.E.B. Methodology: O.W., L.A.S. Formal Analysis: L.A.S, O.W. Investigation: O.W. Resources: S.K., W.G. Data Curation: O.W., CIFASD., S.K. Writing – original draft preparation: O.W., L.A.S. Writing – review and editing: O.W., L.A.S, S.K., W.G., C.E.B., C.G.B. Visualization: O.W., L.A.S. Supervision. C.E.B., C.G.B., L.S. Funding Acquisition. C.E.B., C.G.B. All or part of this work was done in conjunction with the Collaborative Initiative on Fetal Alcohol Spectrum Disorders (CIFASD, https://doi.org/10.5967/ntw9-h991), which is funded by grants from the National Institute on Alcohol Abuse and Alcoholism (NIAAA;). Additional information about CIFASD, including information describing available data, can be found at https://www.cifasd.org. We would a like to thank Dr. Leah Wetherill and Abigail Erickson for support on the assembly of the data dictionary, and Jennifer Galdieri for literature review for the manuscript introduction. ChatGPT 5.2 (OpenAI) was used solely for language editing and text refinement; authors retain full responsibility for the content and conclusions of the manuscript.

## Sources of Funding

O. Weeks was supported by an American Heart Association (AHA) Postdoctoral Fellowship (24POST1192144) and a National Institutes of Health (NIH) training grant awarded to Boston Children’s Hospital (BCH; T32HL007572; Principal Investigator (PI); W.T. Pu). This work was supported by an NIH U01 grant from the NIAAA (U01AA030185). Additional support to the Burns laboratory includes NIH grants R01HL176663 and R01HL171206 and funds from the Boston Children’s Hospital Department of Cardiology. L. Sleeper was supported by funds from the Boston Children’s Hospital Heart Center. S. Khurshid was supported by AHA 23CDA1050571 and NIH K23HL169839. W. Goessling was supported by NIH R01DK135270, R01DK135271, R24OD035402.

## Disclosures

None.

## Supplemental Material

Figure S1 - S3

Table S1 - S5

## Notes

### Competing Interest Statement

The authors have declared no competing interest.

### Author Declarations

The Mass General Brigham institutional review board approved the current study (2017P000752), and informed consent was waived. Mass General Brigham IRB Mass General Brigham 399 Revolution Drive, Suite 710 Somerville, MA 02145 Tel: 857-282-1900 Fax: 857-282-5693

### Summary of Updates

We have added an additional figure outlining the work flow of the cohort study, updated the discussion section, and included a sensitivity analysis for prior statistics.

## Works Cited

1. Popova S, Lange S, Probst C, Parunashvili N, and Rehm J. Prevalence of alcohol consumption during pregnancy and Fetal Alcohol Spectrum Disorders among the general and Aboriginal populations in Canada and the United States. Eur J Méd Genet. 2017;60(1):32–48.

2. Wozniak JR, Riley EP, and Charness ME. Clinical presentation, diagnosis, and management of fetal alcohol spectrum disorder. Lancet Neurology. 2019.

3. May PA, Baete A, Russo J, Elliott AJ, Blankenship J, Kalberg WO, et al. Prevalence and Characteristics of Fetal Alcohol Spectrum Disorders. Pediatrics. 2014;134(5):855–66.

4. Hoyme EH, Kalberg WO, Elliott AJ, Blankenship J, Buckley D, Marais A-S, et al. Updated Clinical Guidelines for Diagnosing Fetal Alcohol Spectrum Disorders. Pediatrics. 2016;138(2):e20154256.

5. Jones KL, Smith DW, and Hanson JW. The Fetal Alcohol Syndrome: Clinical Deliniation Ann Ny Acad Sci. 1976;273:130–7.

6. Jones KL, and W. SD. Recognition of the fetal alcohol syndrome in early infancy. Lancet. 1973;302(7836):999–1001.

7. Burd L, Deal E, Rios R, Adickes E, Wynne J, and Klug MG. Congenital Heart Defects and Fetal Alcohol Spectrum Disorders. Congenit Heart Dis. 2007;2(4):250–5.

8. Adams MM, Mulinare J, and Dooley K. Risk factors for conotruncal cardiac defects in Atlanta. J Am Coll Cardiol. 1989;14(2):432–42.

9. Fung A, Manlhiot C, Naik S, Rosenberg H, Smythe J, Lougheed J, et al. Impact of Prenatal Risk Factors on Congenital Heart Disease in the Current Era. J Am Hear Assoc: Cardiovasc Cerebrovasc Dis. 2013;2(3):e000064.

10. Kurita H, Motoki N, Inaba Y, Misawa Y, Ohira S, Kanai M, et al. Maternal alcohol consumption and risk of offspring with congenital malformation: the Japan Environment and Children’s Study. Pediatr Res. 2021;90(2):479–86.

11. Mills JL, and Graubard BI. Is moderate drinking during pregnancy associated with an increased risk for malformations? Pediatrics. 1987;80(3):309–14.

12. Sun J, Chen X, Chen H, Ma Z, and Zhou J. Maternal Alcohol Consumption before and during Pregnancy and the Risks of Congenital Heart Defects in Offspring: A Systematic Review and Meta-analysis. Congenit Heart Dis. 2015;10(5):E216–E24.

13. Wen Z, Yu D, Zhang W, Fan C, Hu L, Feng Y, et al. Association between alcohol consumption during pregnancy and risks of congenital heart defects in offspring: meta-analysis of epidemiological observational studies. Ital J Pediatr. 2016;42(1):12.

14. Yang J, Qiu H, Qu P, Zhang R, Zeng L, and Yan H. Prenatal Alcohol Exposure and Congenital Heart Defects: A Meta-Analysis. Plos One. 2015;10(6):e0130681.

15. Dorsey AN, Downing KF, Deputy NP, Weber MK, and Howards PP. Prevalence of congenital heart defects among children with and without diagnosed fetal alcohol spectrum disorders, 2016–2022. Drug Alcohol Depend. 2025;274:112790.

16. Weeks O, Gao X, Basu S, Galdieri J, Chen K, Burns CG, and Burns CE. Embryonic alcohol exposure in zebrafish predisposes adults to cardiomyopathy and diastolic dysfunction. Cardiovasc Res. 2024:cvae139.

17. Jonathan CC, Mary Ellen L, and Claire DC. Association Analysis: Fetal Alcohol Spectrum Disorder and Hypertension Status in Children and Adolescents. Alcohol Clin Exp Res. 2019;43:1727–33.

18. Himmelreich M, Lutke, C.J., Hargrove, E.T. In: Begun ALM, M.M. ed. The Routledge Handbook of Social Work and Addictive Behviors. New York: Routledge; 2020:191–215.

19. Attell BK, Snyder AB, Coles C, and Kable J. Comorbidities associated with fetal alcohol spectrum disorders in the United States. Sci Rep. 2025;15(1):29704.

20. Onesimo R, Rose CD, Delogu AB, Battista A, Leoni C, Veltri S, et al. Two case reports of fetal alcohol syndrome: broadening into the spectrum of cardiac disease to personalize and to improve clinical assessment. Ital J Pediatr. 2019;45(1):167.

21. Weeks O, Bossé GD, Oderberg IM, Akle S, Houvras Y, Wrighton PJ, et al. Fetal alcohol spectrum disorder predisposes to metabolic abnormalities in adulthood. J Clin Investigation. 2020.

22. Khurshid S, Reeder C, Harrington LX, Singh P, Sarma G, Friedman SF, et al. Cohort design and natural language processing to reduce bias in electronic health records research. npj Digit Med. 2022;5(1):47.

23. R Core Team R: A Language and Environment for Statistical Computing, Vienna, Austria. https://www.r-project.org/.

24. Burd L, and Popova S. Fetal Alcohol Spectrum Disorders: Fixing Our Aim to Aim for the Fix. Int J Environ Res Public Heal. 2019;16(20):3978.

