## Supplemental Tables and Figures for "Cardiovascular and Cardiometabolic Outcomes in Adults with Fetal Alcohol Spectrum Disorders: A Retrospective Cohort Study"

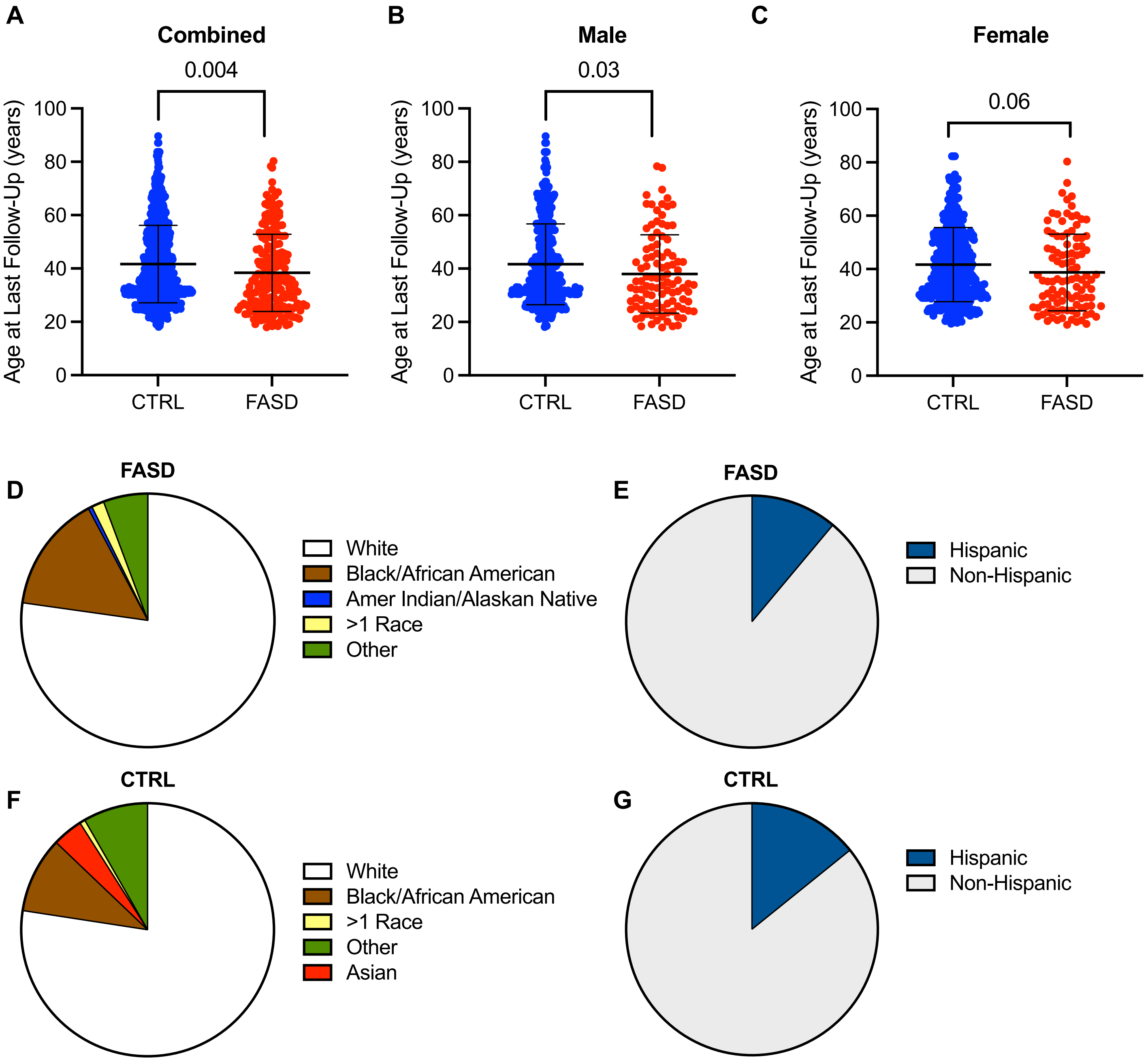

**Supplemental Figure 1. Demographic Characteristics. A – C.** Distribution of age at last follow-up (A); males (B); and females (C). Student t-test. The middle and outer bars represent mean and one standard deviation, respectively. **D – G.** Pie chart depictions of race ($p<0.001$) and ethnicity ($p=0.41$) in FASD (D – E) and CTRL (F – G) cohorts. CTRL=control; FASD=fetal alcohol spectrum disorder.

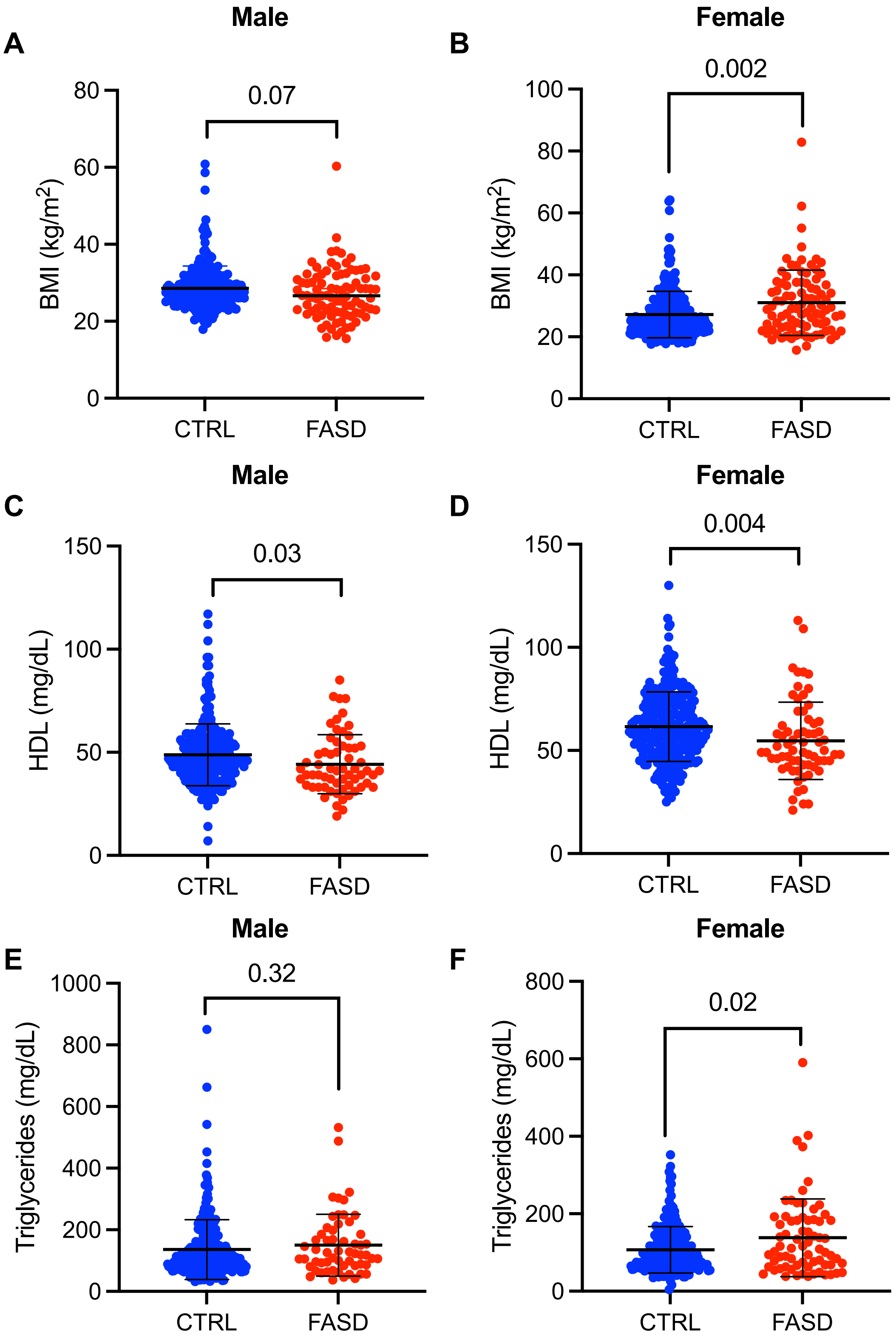

**Supplemental Figure 2. Cardiometabolic parameters at the time of age at last follow-up. A – B.** BMI distribution of male (p = 0.07) and female (p = 0.002) CTRL and FASD cohorts. **C – D.** HDL distribution of male (p = 0.03) and female (p = 0.004) CTRL and FASD cohorts. **E – F.** Distribution of triglyceride level in male (p=0.32) and female (p=0.02) CTRL and FASD cohorts. Student's t-test. The middle and outer bars represent mean and one standard deviation, respectively See Table 2 for age-adjusted mean BMI estimates by sex. CTRL=control; FASD=fetal alcohol spectrum disorder; BMI=body mass index; HDL=high-density lipoprotein.

**Supplemental Table 1. Cardiometabolic Parameters at Age of Last Follow-Up**

| | Unadjusted Mean+SD or n (%) | | |
|---|---|---|---|
| **Parameter** | **FASD** | **Control** | ***P* value** |
| N | 208 | 824 | |
| Body mass index (kg/m$^2$) * | 29.1±9.1 | 27.9±6.8 | 0.09 |
| Weight Category * | | | **0.02** |
| Underweight | 6 (3.3%) | 9 (1.7%) | |
| Normal | 57 (31.5%) | 182 (35.1%) | |
| Overweight | 49 (27.1%) | 183 (35.3%) | |
| Obese | 69 (38.1%) | 145 (27.9%) | |
| LDL cholesterol (mg/dL) † | 99.6±36.4 | 108.5±37.3 | **0.046** |
| HDL cholesterol (mg/dL) ‡ | 49.7±17.5 | 55.1±17.2 | **0.001** |
| HDL cholesterol (< 40 mg/dL) ‡ | 39 (30.2%) | 92 (17.0%) | **0.001** |
| Triglycerides (mg/dL) § | 143.5±100.1 | 122.1±82.3 | **0.03** |
| Hyperlipidemia | 73 (35.1%) | 221 (26.8%) | **0.02** |
| Type 2 diabetes mellitus | 28 (13.5%) | 40 (4.9%) | **<0.001** |
| Multiple cardiometabolic abnormalities ‖ # | 44 (34.9%) | 78 (20.0%) | **0.001** |

[*]Reduced sample size due to missing data. n=181 (FASD); 519 (CTRL)
[†]Reduced sample size due to missing data. n=123 (FASD); 152 (CTRL)
[‡]Reduced sample size due to missing data. n=129 (FASD); 542 (CTRL)
[§]Reduced sample size due to missing data. n=128 (FASD); 487 (CTRL)
[‖]Reduced sample size due to missing data. n=126 (FASD); 390 (CTRL)
[#]Includes 3 factors: overweight/obese; HDL cholesterol < 40 mg/dL; type 2 diabetes mellitus
FASD=Fetal alcohol spectrum disorder; HDL=high-density lipoprotein.

**Supplemental Table 2. Prevalence: Hypertension and Adult Cardiovascular Diseases by Sex.**

| Unadjusted % | | | | | | | | | | |
|---|---|---|---|---|---|---|---|---|---|---|
| | COMBINED | | | MALES | | | FEMALES | | | |
| **Condition** | **FASD** | **Control** | ***P* value** | **FASD** | **Control** | ***P value*** | **FASD** | **Control** | ***P value*** | **Sex x Group Interaction p-value** |
| N | 208 | 824 | | 103 | 406 | | 105 | 418 | | |
| Hypertension | 52 (25.0%) | 185 (22.4%) | 0.46 | 22 (21.4%) | 114 (28.1%) | 0.21 | 30 (28.6%) | 71 (17.0%) | **0.01** | **0.004** |
| Any cardiac condition (excluding HTN) | 50 (24.0%) | 91 (11.0%) | **<0.001** | 24 (23.3%) | 54 (13.3%) | **0.01** | 26 (24.8%) | 37 (8.9%) | **0.001** | 0.17 |
| Conduction defect | 15 (7.2%) | 15 (1.8%) | **<0.001** | 8 (7.8%) | 7 (1.7%) | **0.004** | 7 (6.7%) | 8 (1.9%) | **0.02** | 0.72 |
| Cardiac arrhythmia | 17 (8.2%) | 29 (3.5%) | **0.007** | 7 (6.8%) | 22 (5.4%) | 0.63 | 10 (9.5%) | 7 (1.7%) | **<0.001** | **0.02** |
| Atrial fibrillation | 8 (3.9%) | 18 (2.2%) | 0.21 | 5 (4.9%) | 16 (3.9%) | 0.59 | 3 (2.9%) | 2 (0.5%) | 0.06 | 0.11 |
| Structural heart remodeling | 22 (10.6%) | 43 (5.2%) | **0.007** | 13 (12.6%) | 32 (7.9%) | 0.17 | 9 (8.6%) | 11 (2.6%) | **0.009** | 0.21 |
| Left atrial enlargement | 11 (5.3%) | 28 (3.4%) | 0.22 | 6 (5.8%) | 20 (4.9%) | 0.80 | 5 (4.8%) | 8 (1.9%) | 0.15 | 0.31 |
| Right atrial enlargement | 3 (1.4%) | 4 (0.5%) | 0.15 | 2 (1.9%) | 3 (0.7%) | 0.27 | 1 (1.0%) | 1 (0.2%) | 0.36 | 0.81 |
| LV dilation/hypertrophy | 16 (7.7%) | 28 (3.4%) | **0.01** | 10 (9.7%) | 22 (5.4%) | 0.11 | 6 (5.7%) | 6 (1.4%) | **0.02** | 0.26 |
| RV dilation/hypertrophy | 7 (3.4%) | 9 (1.1%) | **0.03** | 5 (4.9%) | 8 (2.0%) | 0.15 | 2 (1.9%) | 1 (0.2%) | 0.10 | 0.38 |
| Heart Failure (HF) | 12 (5.8%) | 15 (1.8%) | **0.005** | 5 (4.9%) | 12 (3.0%) | 0.36 | 7 (6.7%) | 3 (0.7%) | **<0.001** | **0.04** |
| Systolic HF | 9 (4.3%) | 11 (1.3%) | **0.01** | 6 (5.8%) | 11 (2.7%) | 0.13 | 3 (2.9%) | 0 (0%) | **0.008** | **0.04** |
| Diastolic HF | 7 (3.4%) | 8 (1.0%) | **0.02** | 2 (1.9%) | 4 (1.0%) | 0.35 | 5 (4.8%) | 4 (1.0%) | **0.02** | 0.38 |
| Systolic or Diastolic Dysfunction | 13 (6.3%) | 19 (2.3%) | **0.006** | 6 (5.8%) | 15 (3.7%) | 0.40 | 7 (6.7%) | 4 (1.0%) | **0.002** | **0.05** |
| Myocardial infarction | 12 (5.8%) | 12 (1.5%) | **<0.001** | 7 (6.8%) | 12 (3.0%) | 0.08 | 5 (4.8%) | 0 (0%) | **<0.001** | **0.01** |
| Stroke/Cerebral Vascular Accident | 8 (3.9%) | 6 (0.7%) | **0.002** | 2 (1.9%) | 4 (1.0%) | 0.35 | 6 (5.7%) | 2 (0.5%) | **0.001** | 0.11 |
| Ischemic * | 3 (1.5%) | 6 (0.7%) | 0.39 | 1 (1.0%) | 4 (1.0%) | - | 2 (2.0%) | 2 (0.5%) | 0.17 | 0.32 |
| Hemorrhagic | 1 (0.5%) | 0 (0%) | 0.20 | 1 (1.0%) | 0 (0%) | 0.20 | 0 (0%) | 0 (0%) | - | - |
| Thrombosis or embolism | 9 (4.3%) | 9 (1.1%) | **0.004** | 2 (1.9%) | 5 (1.2%) | 0.63 | 7 (6.7%) | 4 (1.0%) | **0.002** | 0.14 |

* Reduced sample size due to missing data on type of stroke. n=103 (FASD male); 406 (CTRL male); 101 (FASD female); 418 (FASD female).
FASD=Fetal alcohol spectrum disorder; HTN=hypertension; LV=left ventricle; RV=right ventricle; HF=heart failure.

**Supplemental Table 3. Age-Adjusted Prevalence: Hypertension and Adult Cardiovascular Diseases.**

| Age-Adjusted %±SE | | | |
|---|---|---|---|
| **Condition** | **FASD** | **Control** | ***P value*** |
| N | | | |
| Hypertension | 26.2±3.4 | 18.3±1.5 | **0.02** |
| Any cardiac condition (excluding HTN) | 24.2±3.2 | 8.8±1.0 | **<0.001** |
| Conduction defect | 7.0±1.8 | 1.6±0.4 | **<0.001** |
| Cardiac arrhythmia | 7.1±1.8 | 2.4±0.5 | **<0.001** |
| Atrial fibrillation | 2.8±1.1 | 1.2±0.4 | 0.06 |
| Structural heart remodeling | 9.5±2.1 | 3.7±0.7 | **<0.001** |
| Left atrial enlargement | 3.9±1.2 | 1.9±0.5 | 0.05 |
| Right atrial enlargement | 1.0±0.7 | 0.3±0.2 | 0.08 |
| LV dilation/hypertrophy | 6.0±1.7 | 1.9±0.5 | **<0.001** |
| RV dilation/hypertrophy | 3.4±1.3 | 1.1±0.4 | **0.02** |
| Heart Failure (HF) | 4.7±1.5 | 1.2±0.4 | **<0.001** |
| Systolic HF | 3.9±0.01 | 1.0±0.3 | **0.003** |
| Diastolic HF | 1.9±0.9 | 0.4±0.2 | **0.003** |
| Systolic or Diastolic Dysfunction | 4.8±1.5 | 1.3±0.4 | **<0.001** |
| Myocardial infarction | 5.3±0.02 | 1.1±0.4 | **<0.001** |
| Stroke/Cerebral Vascular Accident | 3.9±1.4 | 0.7±0.3 | **0.002** |
| Ischemic * | 1.3±0.8 | 0.6±0.3 | 0.23 |
| Hemorrhagic | 0.1±0.3 | 0 | 0.96 |
| Thrombosis or embolism | 4.2±0.01 | 1.1±0.4 | **0.004** |

* Reduced sample size due to missing data on type of stroke. n=103 (FASD male); 406 (CTRL male); 101 (FASD female); 418 (FASD female).

FASD=Fetal alcohol spectrum disorder; HTN=hypertension; LV=left ventricle; RV=right ventricle; HF=heart failure.

### Supplemental Table 4. Echocardiography Parameters

| | Unadjusted Mean±SD | | | | | | | | | | | | | |
|---|---|---|---|---|---|---|---|---|---|---|---|---|---|---|
| | COMBINED | | | | | MALES | | | | FEMALES | | | | |
| **Parameter** | **N** | **FASD** | **N** | **Control** | ***P* value** | **N** | **FASD** | **N** | **Control** | **N** | **FASD** | **N** | **Control** | **Sex x Group Interaction p-value** |
| LV ejection fraction (%) | 52 | 59.6±13.2 | 120 | 63.2±8.8 | 0.07 | 23 | 58.9±17.5 | 68 | 61.2±10.2 | 29 | 60.1±8.8 | 52 | 65.9±5.5 | 0.32 |
| LV end-diastolic internal dimension | 41 | 45.8±8.4 | 103 | 46.4±5.7 | 0.65 | 18 | 46.5±11.3 | 58 | 47.8±5.9 | 23 | 45.2±5.5 | 45 | 44.7±4.9 | 0.46 |
| LV end-systolic internal dimension | 40 | 32.7±9.6 | 98 | 30.2±5.4 | 0.13 | 17 | 34.0±12.1 | 52 | 31.3±6.0 | 23 | 31.8±7.4 | 46 | 29.0±4.41 | 0.98 |
| Interventricular septum (IVS) | 39 | 9.3±2.6 | 94 | 10.0±2.1 | 0.10 | 19 | 9.7±3.1 | 52 | 10.6±2.1 | 20 | 8.9±2.1 | 36 | 9.2±1.9 | 0.45 |
| Posterior wall thickness (PWT) | 34 | 9.2±2.8 | 88 | 9.9±2.3 | 0.21 | 19 | 9.5±3.1 | 52 | 10.3±2.6 | 15 | 8.8±2.6 | 36 | 9.2±1.6 | 0.71 |
| | **Age-Adjusted Mean±SE** | | | | | | | | | | | | | |
| | **COMBINED** | | | | | **MALES** | | | | **FEMALES** | | | | |
| **Parameter** | **N** | **FASD** | **N** | **Control** | ***P* value** | **N** | **FASD** | **N** | **Control** | **N** | **FASD** | **N** | **Control** | **Sex x Group Interaction p-value** |
| LV ejection fraction (%) | 52 | 58.9±1.4 | 120 | 64.0±0.9 | **0.004** | 23 | 57.3±2.2 | 68 | 62.4±1.2 | 29 | 60.1±1.9 | 52 | 65.9±1.4 | 0.85 |
| LV end-diastolic internal dimension | 41 | 46.0±1.1 | 103 | 46.4±0.7 | 0.72 | 18 | 47.0±1.6 | 58 | 47.8±0.9 | 23 | 45.2±1.4 | 45 | 44.7±1.0 | 0.64 |
| LV end-systolic internal dimension | 40 | 32.9±1.1 | 98 | 30.2±0.7 | **0.046** | 17 | 34.8±1.8 | 52 | 31.4±1.0 | 23 | 31.6±1.5 | 46 | 28.9±1.0 | 0.77 |
| Interventricular septum (IVS) | 39 | 9.7±0.3 | 94 | 9.9±0.2 | 0.59 | 19 | 10.4±0.5 | 52 | 10.5±0.3 | 20 | 9.1±0.4 | 36 | 9.2±0.3 | 0.64 |
| Posterior wall thickness (PWT) | 34 | 9.6±0.4 | 88 | 9.8±0.2 | 0.62 | 19 | 10.2±0.6 | 52 | 10.2±0.3 | 15 | 8.9±0.6 | 36 | 9.3±0.4 | 0.73 |

FASD=Fetal alcohol spectrum disorder; LV=left ventricle.

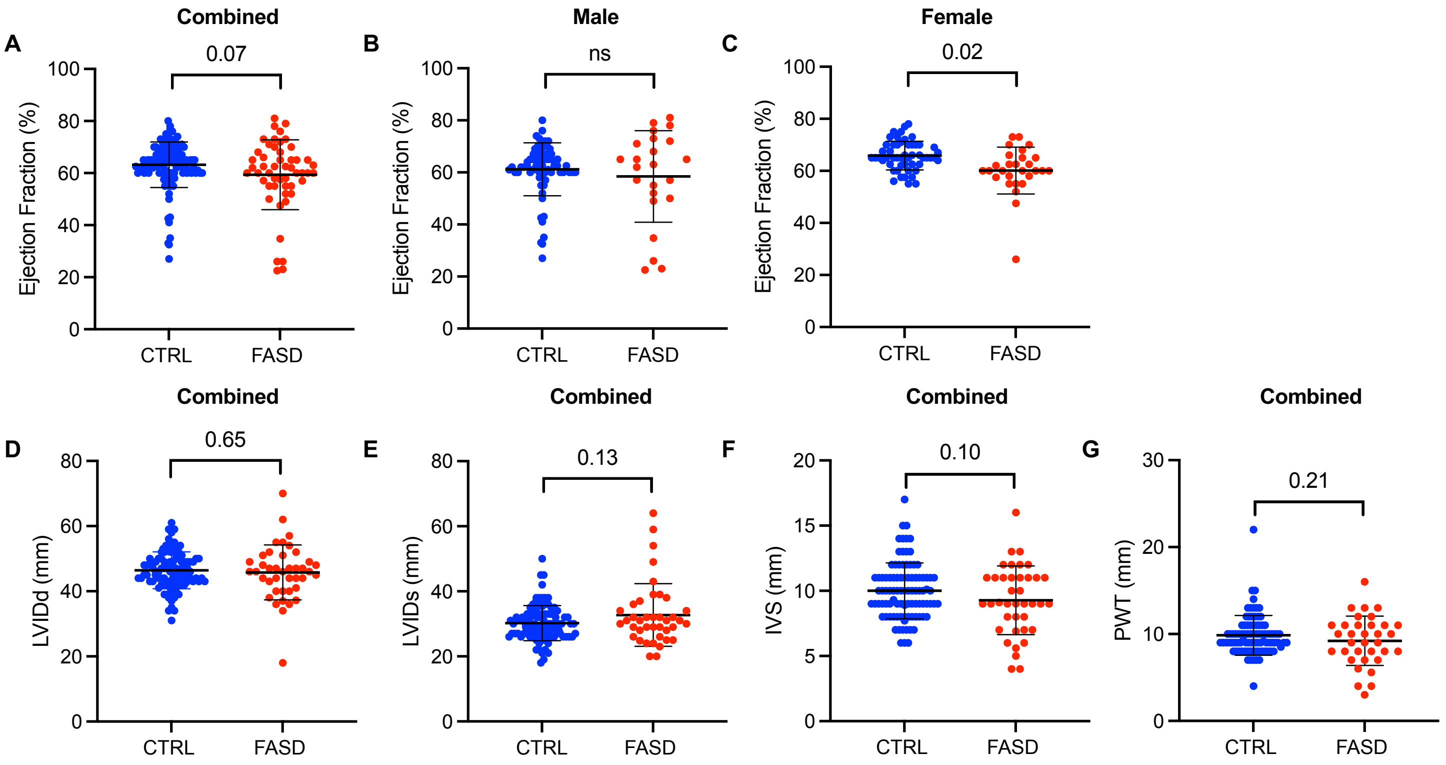

**Supplemental Figure 3. Echocardiography Parameters (unadjusted). A – C.** Distribution of left ventricular ejection fraction (%) in the combined cohort (p = 0.07), males (p = 0.35), and females (p = 0.02) from the FASD and CTRL cohorts. **D – G.** Distribution of left ventricular internal dimension during diastole (LVIDd; p = 0.65), left ventricular internal dimension during systole (LVIDs; p = 0.13), intraventricular septum thickness (IVS; p = 0.1), and posterior wall thickness (PWT; p = 0.21) from echocardiography reports in FASD and CTRL cohorts. The maximum sample size across all measures was 52 for FASD and 120 for CTRL. Student's t-test. The middle and outer bars represent mean and one standard deviation, respectively. FASD=fetal alcohol spectrum disorder; CTRL= control.

**Supplemental Table 5. Mean Age at Last Follow-Up by CHD Status in the FASD Cohort**

| CHD | N | Mean | Median | IQR | Minimum | Maximum |
|---|---|---|---|---|---|---|
| **NO** | 195 | 38.4±14.7 | 35.3 | (26.0, 48.0) | 18.0 | 80.3 |
| **YES** | 13 | 38.0±11.9 | 36.2 | (29.0, 41.9) | 22.9 | 58.3 |

CHD=congenital heart defect.

**Supplemental Table 6 (Related to Table 6): Cardiovascular Condition (Excluding Hypertension) or Heart Remodeling Within Subgroup, FASD and Control**

| % with Cardiovascular Condition (Excluding Hypertension) | | | |
|---|---|---|---|
| **Condition** | **FASD** | **Control** | **p-value** |
| Hyperlipidemia | 23% (17/73) | 19% (43/221) | 0.50 |
| No hyperlipidemia | 24% (33/135) | 8% (48/603) | <0.001 |
| Multiple cardiometabolic abnormalities* | 27% (12/44) | 24% (19/78) | 0.83 |
| No multiple cardiometabolic abnormalities* | 28% (23/82) | 12% (36/312) | <0.001 |
| **% with Structural Heart Remodeling** | | | |
| **Condition** | **FASD** | **Control** | **p-value** |
| Hyperlipidemia | 11% (8/73) | 10% (22/221) | 0.82 |
| No hyperlipidemia | 10% (14/135) | 3% (21/603) | 0.002 |
| Multiple cardiometabolic abnormalities* | 16% (7/44) | 19% (15/78) | 0.81 |
| No multiple cardiometabolic abnormalities* | 11% (9/82) | 3% (10/312) | 0.008 |

*Includes 3 factors: overweight/obese; HDL cholesterol<40; type 2 diabetes mellitus
FASD=fetal alcohol spectrum disorder.
